# Polygenic risk score portability for common diseases across genetically diverse populations

**DOI:** 10.1101/2024.06.13.24308905

**Authors:** Sonia Moreno-Grau, Manvi Vernekar, Arturo Lopez-Pineda, Daniel Mas-Montserrat, Miriam Barrabes-Torrella, Consuelo D. Quinto-Cortés, Babak Moatamed, Ming Ta ’Michael’ Lee, Zhenning Yu, Kensuke Numakura, Yuta Matsuda, Jeffrey D. Wall, Alexander Ioannidis, Nicholas Katsanis, Tomohiro Takano, Carlos D Bustamante

**Author notes:** Corresponding author. **Address correspondence to: Dr. Carlos D. Bustamante** Galatea Bio, Inc. 14350 Commerce Way, Miami Lakes, Florida 33016, United States, **Mr. Tomohiro Takano** USA: Genomelink, Inc. 2150 Shattuck Avenue, Berkeley, CA 94704 Japan: Awakens Japan K.K. (Japanese subsidiary of Genomelink, Inc.) 2-11-3, Meguro, Meguro-ku, Tokyo, Japan 1530063.

## Abstract

**Background:** Polygenic risk scores (PRS) derived from European individuals have reduced portability across global populations, limiting their clinical implementation at worldwide scale. Here, we investigate the performance of a wide range of PRS models across four ancestry groups (Africans, Europeans, East Asians, and South Asians) for 14 conditions of high-medical interest.

**Methods:** To select the best-performing model per trait, we first compared PRS performances for publicly available scores, and constructed new models using different methods (LDpred2, PRS-CSx and SNPnet). We used 285K European individuals from the UK Biobank (UKBB) for training and 18K, including diverse ancestries, for testing. We then evaluated PRS portability for the best models in Europeans and compared their accuracies with respect to the best PRS per ancestry. Finally, we validated the selected PRS models using an independent set of 8,417 individuals from Biobank of the Americas-Genomelink (BBofA-GL); and performed a PRS-Phewas.

**Results:** We confirmed a decay in PRS performances relative to Europeans when the evaluation was conducted using the best-PRS model for Europeans (51.3% for South Asians, 46.6% for East Asians and 39.4% for Africans). We observed an improvement in the PRS performances when specifically selecting ancestry specific PRS models (phenotype variance increase: 1.62 for Africans, 1.40 for South Asians and 0.96 for East Asians). Additionally, when we selected the optimal model conditional on ancestry for CAD, HDL-C and LDL-C, hypertension, hypothyroidism and T2D, PRS performance for studied populations was more comparable to what was observed in Europeans. Finally, we were able to independently validate tested models for Europeans, and conducted a PRS-Phewas, identifying cross-trait interplay between cardiometabolic conditions, and between immune-mediated components.

**Conclusion:** Our work comprehensively evaluated PRS accuracy across a wide range of phenotypes, reducing the uncertainty with respect to which PRS model to choose and in which ancestry group. This evaluation has let us identify specific conditions where implementing risk-prioritization strategies could have practical utility across diverse ancestral groups, contributing to democratizing the implementation of PRS.

## 1. Background

Polygenic risk scores (PRS) predict complex disease susceptibility and traits based on genetic data [1]. PRSs have been identified as a promising tool to be implemented from genomic medicine to population-based screenings [2]. However, most of PRS are impacted by a lack of portability, i.e. the variation in predictive performance metrics across populations, which limits their utility.

The portability of PRS depends on genetic similarity between training and target cohorts. Several studies have demonstrated a decay in PRS accuracy with genetic distance from the training set, at continental and subcontinental level [1][3][4][5][6]. Therefore, most European-derived PRS are not likely to predict as well in individuals of other ancestries. The underlying reasons explaining these disparities are commonly attributed to differences in causal variants, linkage disequilibrium (LD) patterns, allele frequencies, and effect sizes across populations. Recent studies have indicated that using PRS derived from multi-ancestry data increases the performance in diverse populations [7]. Despite that, lack of precise PRS models hinders risk stratification and leads to exacerbation of health disparities in implementation of the PRSs across different societies [1].

PRS portability might be improved by reducing the genetic distance from target to training data, which might be promoted by increasing population diversity in large biobanks. According to the GWAS catalog on January 2024, ∼78% of the individuals included were of European ancestry, followed by ∼11% of Asian ancestry, and ∼4% from other minorities (i.e. African, Hispanic/Latino, middle Eastern, Native American and Oceanian ancestries) (https://www.ebi.ac.uk/gwas/docs/ancestry-data). Although active recruitment of non-European individuals in genetic studies is greatly increasing [8][9][10], large biobanks and cohorts are still far from proportionally representing global populations.

Although generalizing a single PRS model useful for all populations supposes the gold standard, it is becoming more established that optimal PRS methods are context specific [11]. Nowadays, a large number of PRS per trait have been developed with different methodologies, scarcely validated, and mostly derived from European individuals. Scarce information is available about their comparative performances in a harmonized scenario across populations. Altogether, it complicates the decision-making with respect to which is the most optimal PRS model for a specific condition and ancestry to be implemented at scale.

Here, we investigate the variability of a wide range of PRS models across four ancestry groups (Africans, Europeans, East Asians and South Asians) for 14 conditions of high-medical interest (i.e. cardiometabolic, digestive, and inflammatory conditions, lipid, and Vitamin D levels). With that objective, we first selected the most optimal PRS model for Europeans from a set of 166 PRS available from PGS Catalog [12] or de novo constructed using three different PRS methods (LDpred2 [13] [14], PRS-CSx [15] and SNPnet [16]) in the UKBB data. Then, to test whether the portability problem might be reduced when choosing the best-performing model in each ancestral group, we compare PRS performances between the best PRS for Europeans and the best PRS per ancestry. Finally, we validated the optimal PRS models per condition in a set of European individuals from the Biobank of the Americas – GenomeLink (BBofA-GL) and performed ancestry-specific PRS-Phewas per condition.

## 2. Methods

### Datasets and Phenotypes

#### UKBB cohort

UKBB is a prospective cohort collected from multiple sites across the United Kingdom, which includes over 500,000 participants. Information on genotyping and quality control has previously been described ^18^. UK Biobank received ethical approval from the NHS National Research Ethics Service North West (11/NW/0382). This research has been conducted under Application Number 89,006.

UKBB phenotypes were extracted through ICD10 codes (See Supplementary Table 1). We explored the following conditions: cardiometabolic (atrial fibrillation [AF], coronary artery disease [CAD], hypertension, type 1 diabetes [T1D], type 2 diabetes [T2D] and hypothyroidism), digestive (celiac disease [CD], hemorrhoids and ulcerative colitis [UC]), and inflammatory conditions (gout), lipid levels (high-density lipoproteins cholesterol [HDL-C]; low-density lipoproteins cholesterol [LDL-C] and triglycerides [TG]), and Vitamin D levels. Briefly, we excluded all individuals with T2D diagnoses from T1D study composition (both case, and controls), and likewise the opposite. Lipid and Vitamin D levels were dichotomized (case vs control) according to a specific cut-off, i.e. LDL > 190 mg/dL or 4.9 mmol/L; HDL > 60 mg/dL or 1.5 mmol/L; Triglycerides > 200 mg/dL or 2.3 mmol/L; and Vitamin D > 30 nmol/L or 12 ng/mL. Control groups per each condition were defined as participants not diagnosed with the tested condition. The final number of cases and the sample demographics for both the training and testing sets are described in the Supplementary Tables 2 and 3.

#### The Biobank of the Americas-Genomelink (BBofA-GL)

The BBofA-GL cohort includes participants drawn from the customer base of Genomelink (genomelink.io), which offers a direct-to-consumer genetic traits platform with more than 500,000 users globally. All participants included in the analyses provided informed consent and answered surveys online according to our human subjects protocol, which was reviewed and approved by WCG IRB (https://www.wcgirb.com/) under IRB tracking ID 2020-1332.

BbofA-GL phenotypes were extracted from self-administered questionnaires. The online survey included specific questions about the 14 tested phenotypes and questions about age, sex, weight, and height. Supplementary Table 4 shows the online questionnaire. Data were collected over a period of six months, from August 2022 to February 2023. Only the initial response of each participant was included in the study if genotype information was available. Case-control groups were created as previously described. We observed an increase in the % of women collected by BbofA-GL, which we explain by using a self-reported questionnaire.

### Genotype data preparation and Quality control

#### UKBB cohort: Training and testing samples

We used the UK Biobank dataset (directly genotyped data from release version 2, and the imputed dataset v3) as part of our training and testing samples. Briefly, we removed individuals from our analyses based on the following criteria reported by the UK Biobank in the sample QC file, “ukb_sqc.txt”: 1) individuals showing putative sex chromosome aneuploidy; 2) marked as outliers for heterozygosity and missing rates; 3) presenting an excess of relatives in the dataset (>10); and we kept individuals used in the principal component analysis (PCA). This process generated a set of 406,000 unrelated individuals.

Then, with the objective of estimating genetic ancestry, we performed a principal component analysis (PCA) using the directly genotyped UKBB version. This analysis was restricted to variants particularly used by the UK Biobank team for internal PCA, extracted from the variant QC file “ukb_snp_qc.txt”. PCA was conducted after merging the dataset with the 1000G reference panel, using –pca option of Plink2 software [17] (https://www.cog-genomics.org/plink/2.0/). We then combined the information derived from self-reported ethnicity (UK Biobank field ID: 21000) and principal component analysis (thresholds on PC1 and PC2), to generate the 4 subpopulations for this study: European (n = 381,199 individuals), African (8,111), East Asian (2,365), and South Asian (SAS, 6,596). The SAS group self-identified as Pakistani, Bangladeshi, Indian or Asian_or_Asian_British. Among them, we used 75% of the directly genotyped data for PRS training and 25% of the imputed data for further PRS evaluation.

In the UK Biobank imputed version (v3), we only used bi-allelic variants with imputation information score ≥ 0.7, MAF ≥ 0.01, and variant call rate ≥ 97% (N = 8,962,898).

#### BbofA-GL: Validation Sample

This cohort includes data from nine independent genotyping arrays. Genotype-level data for each array were processed by applying identical quality control and imputation procedures, as previously described [18]. Briefly, variants with a call rate of < 97% and palindromic markers (A/T, G/C, MAF > 0.4) were excluded. We performed an exact test for Hardy–Weinberg equilibrium for individuals of the largest ancestral group (p < 1 × 10^−12^, globally). Individual quality control (QC) includes matching between gender identification and chromosomal sex (when it is possible), and no excess ancestry-adjusted heterozygosity. Samples genetically related to other individuals in the cohort and duplicates were detected and removed using the King algorithm (–make-king, king estimate > 0.177; Plink2 [17]). PCA was performed to identify global ancestry per individual using 1000 Genomes Project data as reference populations (See Supplementary Figure 2). Further information about the number of markers and samples per genotyping array pre- and post-QC is available in Supplementary Table 5. Imputation was carried out using 1000 Genomes Project data as a reference panel with Beagle [19]. Next, we generated a merged dataset combining imputed genotypes (MAF > 0.01; imputation quality R2 > 0.30) from available datasets. Imputed makers with call rate > 0.97 in the merged data were selected for downstream analysis (N = 9,605,881). The final sample size for this dataset was 8,394 individuals of European ancestry with all available phenotypes.

### Polygenic Risk Score Calculation

We used the --score function from Plink2 [19] to calculate PRS. Briefly, for each individual j, the PRS score takes the form PRSj=∑βiGij; where βi is a weight equal to the log odds ratios for the variant *i*, and Gij is the number of risk alleles (2,1,0) of variant *i* in subject *j.* The weights for each variant were obtained from: a) publicly available models; and b) new models trained using three different PRS methods (LDpred2, PRS-CSx and SNPnet). We specifically followed previous recommendations ^8^ to ensure the coding allele in the target data was the effective allele reported in the summary statistic. PRS-scores were standardized to PRS Z-scores for downstream analyses.

#### PRS Models publicly available

We downloaded a total of 402 PRS models for 14 phenotypes (Supplementary Table 6) from the PGS-Catalog database [12] (https://www.pgscatalog.org/) on April 27, 2023: 14 for AF, 10 for

CD, 35 for CAD, 9 for gout, 3 for hemorrhoids, 33 for HDL, 50 for hypertension, 15 for hypothyroidism, 55 for LDL, 34 for TG, 9 for T1D, and 88 for T2D, 3 for UC and 44 for vitamin D. With the goals of generating automatic downloads and harmonizing data, we used custom versions of the “d*owlnoad_scorefiles.py*” and the “*combine_scorefile.py*” scripts from PGS Catalog Utilities (https://pypi.org/project/pgscatalog-utils/). The studied PRS models were downloaded in GRCh37 genome assembly. To avoid overfitting of the results, we excluded from the European analysis all the downloaded PGS-Catalog models using GWAS summary statistics derived from the UKBB European set or trained with them. In the case of the analysis conducted in non-European individuals, we just limited the number of available scores to those specifically using summary statistics or training with the UKBB non-European set. This lets us have a total of 97 and 400 publicly available models available for testing in the UKBB sample, respectively.

#### PRS Model Training

New PRS models were trained on 285,900 European individuals from UKBB, when we used LDpred2 and SNPnet. When GWAS summary statistics were needed (LDpred2 and PRS-CSx) we downloaded available summary statistics from Finngen [20], version DF9 (https://r9.finngen.fi/), and from Biobank of Japan [21] (https://pheweb.jp/) for the studied phenotypes. Software conditions are herewith detailed:

##### Batch screening iterative lasso (BASIL, or SNPnet in its R implementation) [16]

SNPnet applies a multiple regression method that takes advantage of a LASSO solver and optimizes for datasets that are too big to fit into the memory. BASIL identifies the set of SNPs that most efficiently discriminate between cases and controls. The training was performed with the following parameters: n lambda = 200; alpha=1; ncores=20. Additional parameters were set up to the default.

##### PRS Continuous Shrinkage (PRS-CSx) [15]

PRS-CSx places a continuous shrinkage (CS) prior on SNP effect sizes, which is robust to varying genetic architectures. Here, we applied the default “auto” version of PRS-CSx that obtains weights through the Gibbs sampling algorithm, using the specific reference panels per specific ancestry (i.e., European samples from the 1000 Genomes Project (“EUR reference”) for individuals of European ancestry). To construct multi-ancestry models, we used the “*meta*” flag, to get the combined SNP effect sizes across populations using an inverse-variance-weighted meta-analysis of the population-specific posterior effect size estimates.

##### LDpred2 (bigsnpr) [14]

Ldpred [13] is a method that infers the posterior mean effect size of each marker by using a prior on effect sizes and LD information from an external reference panel (the default value is from HapMap 3). A Gibbs sampler is used to estimate the residualized marginal effect for each SNP. In this work, we used LDpred2, which extends LDpred by auto-tuning the best-performing hyper-parameters within the inner loop of the algorithm. We used SNPs that are overlapped in HapMap 3 variants and UKBB genotypes (Supplementary Table 7).

### Statistical Analysis - Evaluation of Model Performance

To assess the performance of the PRS models, we evaluated: i) area under curve (AUC); ii) percentage of the variance explained by the PRS (%Variance); iii) effect and significance of the PRS after multiple testing correction (Bonferroni correction); and iv) case enrichment across PRS percentiles; using individuals that were not included in the training sets from the UKBB and the BBofA-GL datasets. AUC per each model was estimated using “predict” and “roc” functions from the pROC package in R software for models including a) covariables only and b) a full model with covariables and the PRS. The included covariates in the analysis were: age, sex and the first four principal components (PCs) resulting from ancestry analysis. Array type was also used as a covariate to control the impact of the 9 different array types in the BBofA-GL dataset. The %Variance explained by the PRS was calculated as the difference between the Full model and the covariates only. The PRS effect on each specific phenotype was estimated from a generalized linear model (glm) for a binomial outcome for: a) an unadjusted model (PRS only), for the full model (PRS and covariables); and c) for the full model also including BMI, using R. To account for multiple testing, we used a Bonferroni threshold of *p* < 2.95 × 10^−4^ (0.05/166) for the results of the analysis with European ancestry individuals and *p* < 1.24 × 10^−4^ (0.05/402) for all models. Finally, disease risk was estimated between intermediate PRS percentiles (40-60%) and PRS percentile (0-10%, 10-20%, 20-30%, 30-40%, 60-70%, 70-80%, 80-90%, and 90-100%) by applying a *glm* as previously described. Phenotypes with number of cases < 25, were excluded from downstream analysis. We used the Ggplot package from R to plot results.

#### PRS-Phewas

PRS-phenome-wide association analyses (PRS-Phewas) for each selected PRS were conducted in the UK Biobank testing sample across different ancestries. All the phenotypes studied in this analysis were binary. The association between the 14 PRS scores and each phenotype, excluding the PRS-condition, was assessed by applying a glm for a binary outcome, for the previously described models, the PRS model only, the full model and the full model considering BMI. This study was performed using R. All estimates correspond to 1 SD change of the PRS. A conservative Bonferroni correction for multiple testing that assumes uncorrelated traits yields a p-value threshold of p<2.74×10^-4^ (0.05/182). We used Ggplot in R to plot results.

## 3. Results

With the objective of evaluating PRS portability across different ancestries and comparing best-performing PRS model in each ancestral group, we have tested publicly available PRS models and constructed new models using three different PRS methods (LDpred2, PRS-CSx and SNPnet) for 14 high-interest medical conditions in two independent cohorts including individuals from different ancestries (see flowchart in **Figure 1**). These scores were trained within 285,000 European individuals and tested in 6,595 individuals of African ancestry, 2,473 of East Asian ancestry, 95,300 of European ancestry, and 8,111 of South Asian ancestry from the UK Biobank (See principal component analysis for ancestry in Supplementary Figure 1). We then validated them in 8,417 European individuals from the BbofA-GL cohort (See Supplementary Figure 2). Further description about the demographic information for UKBB and BbofA-GL can be found in Supplementary Tables 2 and 3.

**Figure 1.**
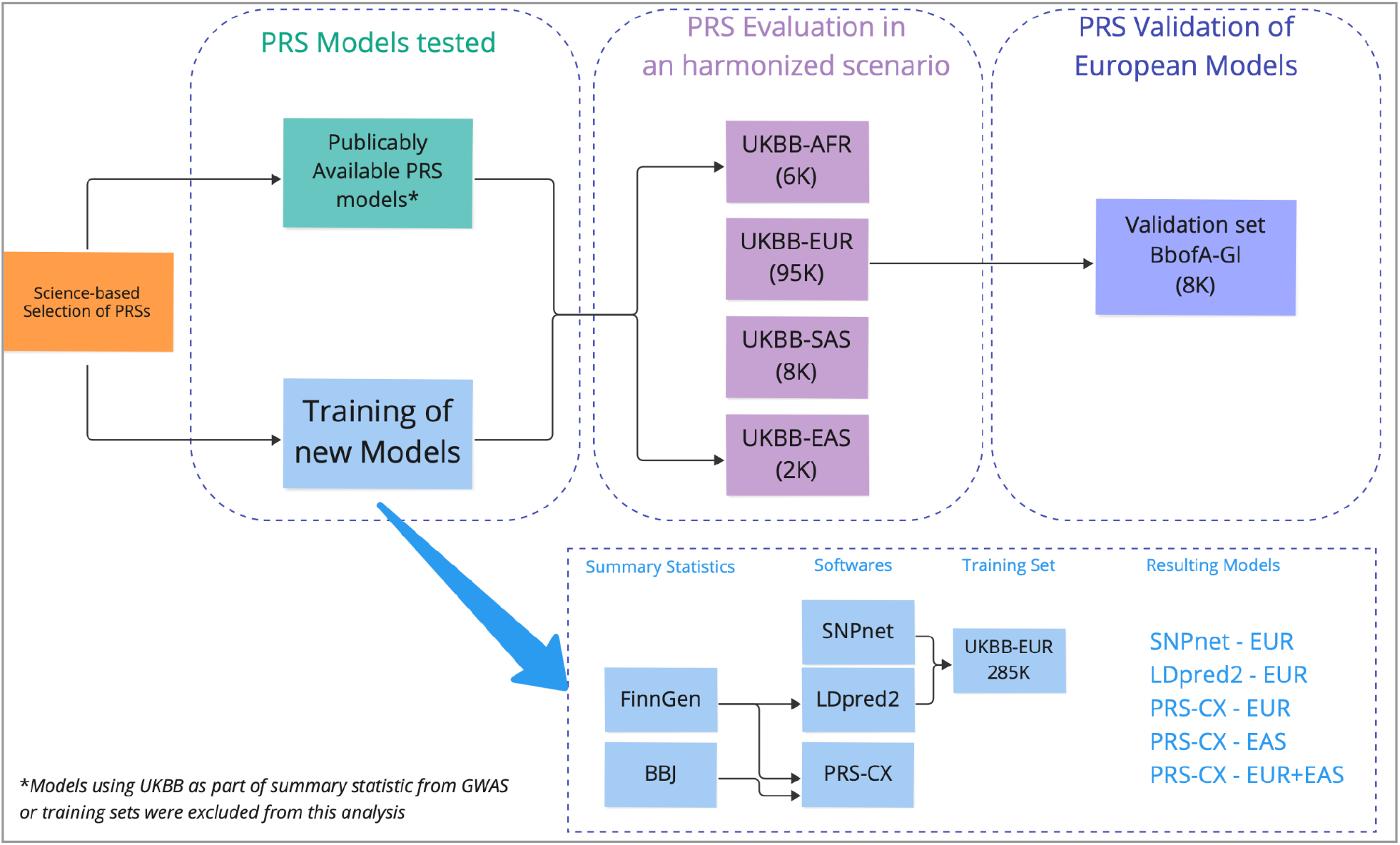
Flow chart of the strategy applied in this study.

### Performance and validation of the optimal PRS models in European-ancestry cohorts

To select the best-performing model per condition, i.e., that exhibiting the highest % of variance explaining the phenotype; we assessed a total of 166 PRS models, including both publicly available and de-novo trained ones, for cardiac, metabolic, digestive, and inflammatory conditions, as well as lipid and vitamin D lab levels. A total of 105 PRSs were significantly associated with their corresponding phenotype (p < 2.74 x 10^-4^) and showed the expected effect direction in the UKBB European set (Supplementary Table 8). The best-performing models per condition were able to explain between 1% for hemorrhoids to ∼22% for celiac disease of disease heritability. We observed effects doubling the risk of developing the condition for UC, CD, T1D, and LDL-C in the testing sample (**Table 1**). UC was the condition showing the strongest effect, with an odds ratio per standard deviation increase (OR/SD) of 3.17 (95% confidence interval (CI) 2.97–3.38) in a model adjusted for age, sex, and principal components for genetic ancestry. The best-performing PRS were then validated in the BbofA-GL cohort. The selected models were significantly replicated in this independent cohort for all the phenotypes, except for HDL and UC (**Table 1**).

**Table 1.**
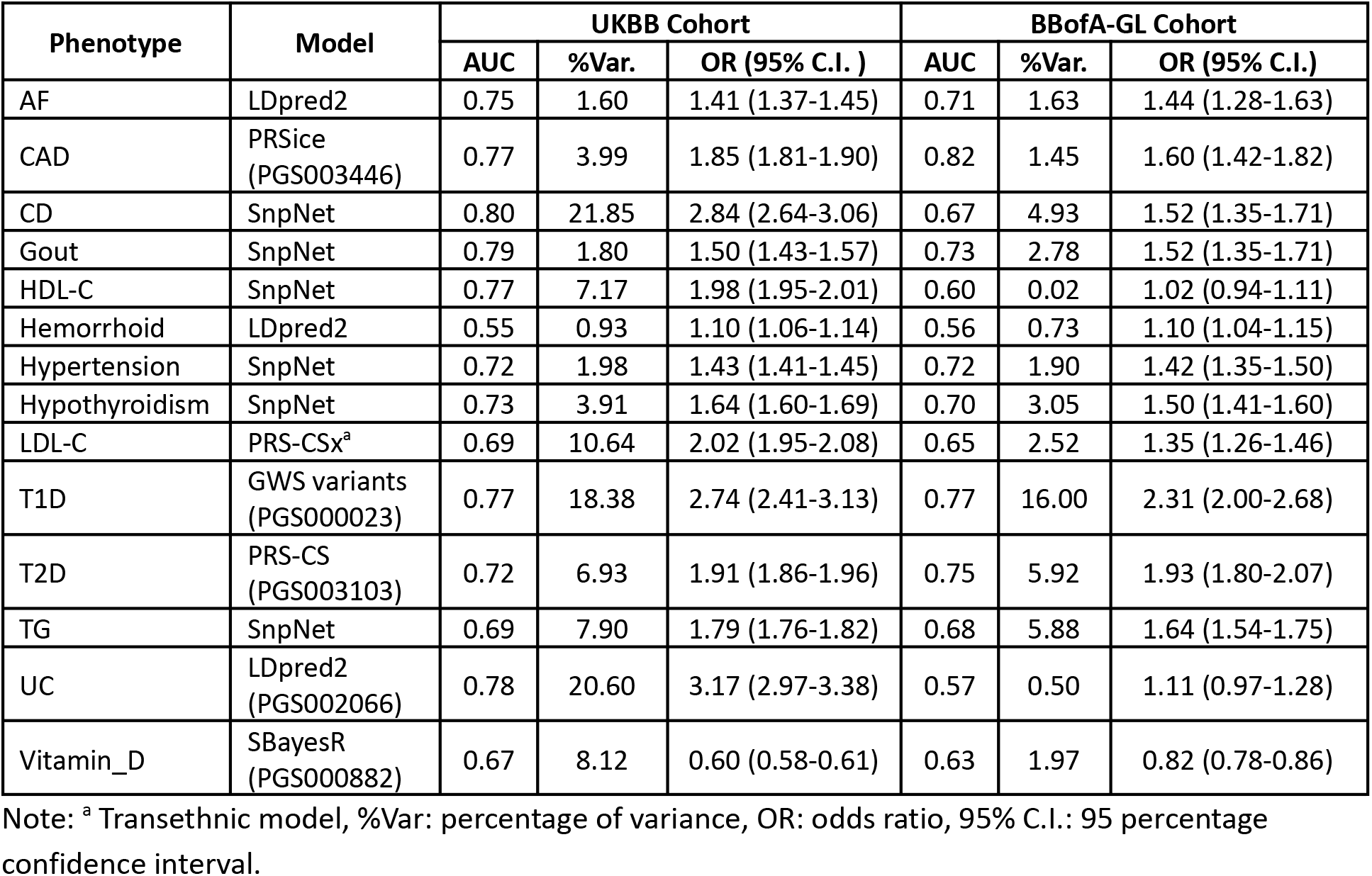
Best-performing polygenic risk score for 14 medical conditions in Europeans.

To estimate what proportion of the population may benefit from PRS assessment, we evaluated case enrichment in the top decile compared to the two central deciles (40-60%). We observed that the top decile was significantly associated with T1D, T2D, CAD, hypertension, hypothyroidism, CD, TG measurement, gout, and Vitamin D, in both the UKBB sample and the BbofA-GL cohort (**Figure 2**, and Supplementary Table 9). Among these conditions, individuals in the top decile had at least two-fold increased risk for T1D, T2D, hypothyroidism, CAD, TG measurement and CD in both datasets (Supplementary Figure 3). When we studied this enrichment exclusively in the UKBB we found at least 2-fold increased risk in an additional four conditions. This disparity could be caused by the different phenotyping strategies used in both datasets, as UKBB data uses ICD10 phecodes and BbofA-GL self-reported questionnaires.

**Figure 2.**
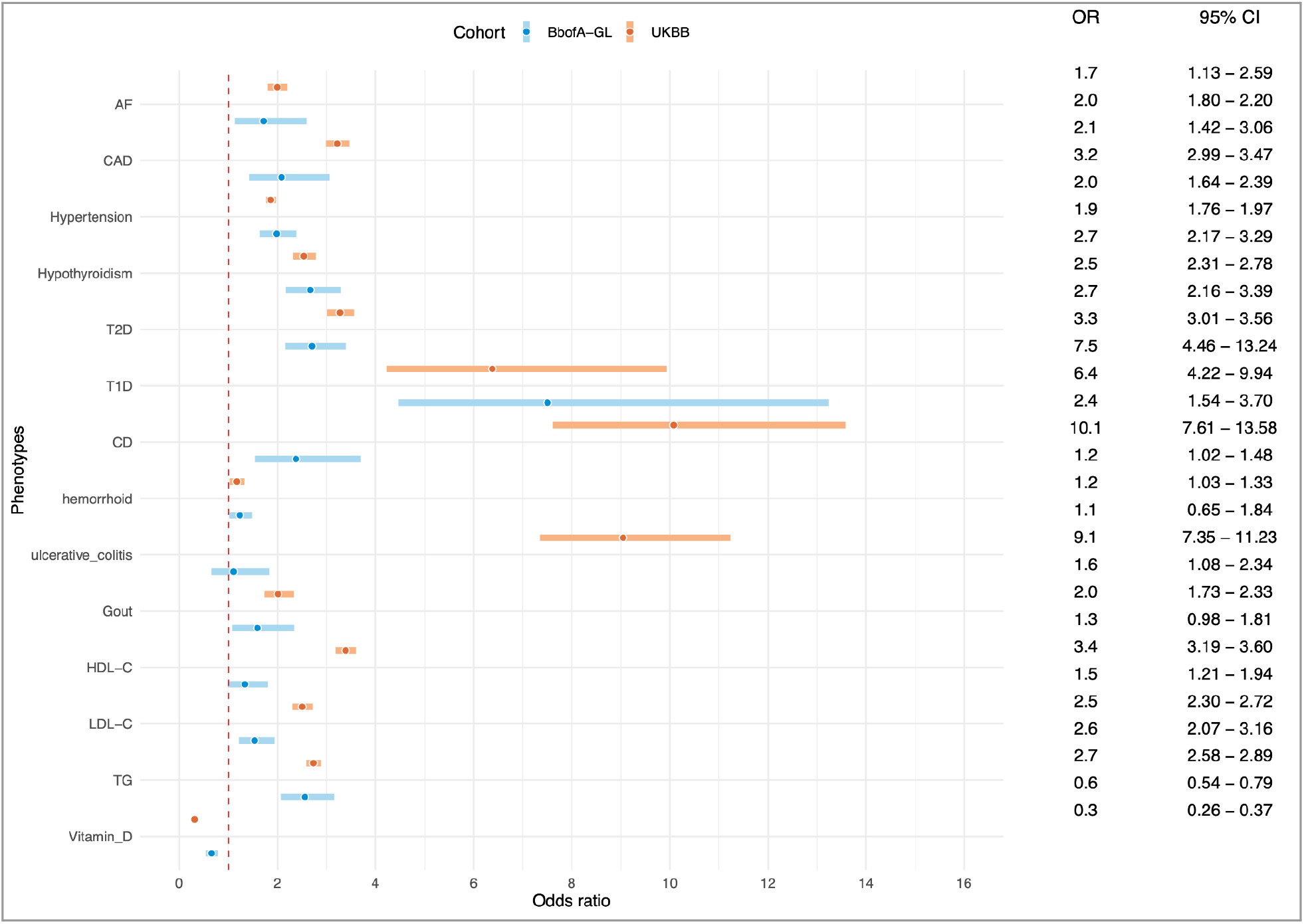
Effect of the 90^th^ PRS percentile compared to the intermediate percentile for fourteen medical conditions in the European set of individuals from the UKBB and BBofA.

### PRS accuracies in groups with non-European ancestries

We proceed to evaluate the portability of the optimal PRS models in Europeans across other populations. We observed a decay in the percentage of variance explained for the European-selected PRS models in non-Europeans (Supplementary Figure 4). The relative predictive performance for the averaged 14 phenotypes compared to prediction performance in Europeans was 51.3% for South Asians, 46.6% for East Asians and 39.4% for Africans.

Considering these results, we modified our approach to select the best-performing model for each condition and ancestral group. We observed a subtle increase in the phenotypic variance explained by the PRS with respect to the observed in the European selected models of 1.62 for Africans, 1.40 for South Asians and 0.96 for East Asians (Table 2). All the scores were significantly associated (p < 1.24 x 10^-4^) with their respective conditions, except for: hemorrhoids in all the ancestries; and AF, hypothyroidism and gout for East Asian ancestry individuals, and Vitamin D in both Asian groups, EAS, and SAS (Table 2). Further information about the performance of the whole set of tested models is provided in Supplementary Table 10.

**Table 2.**
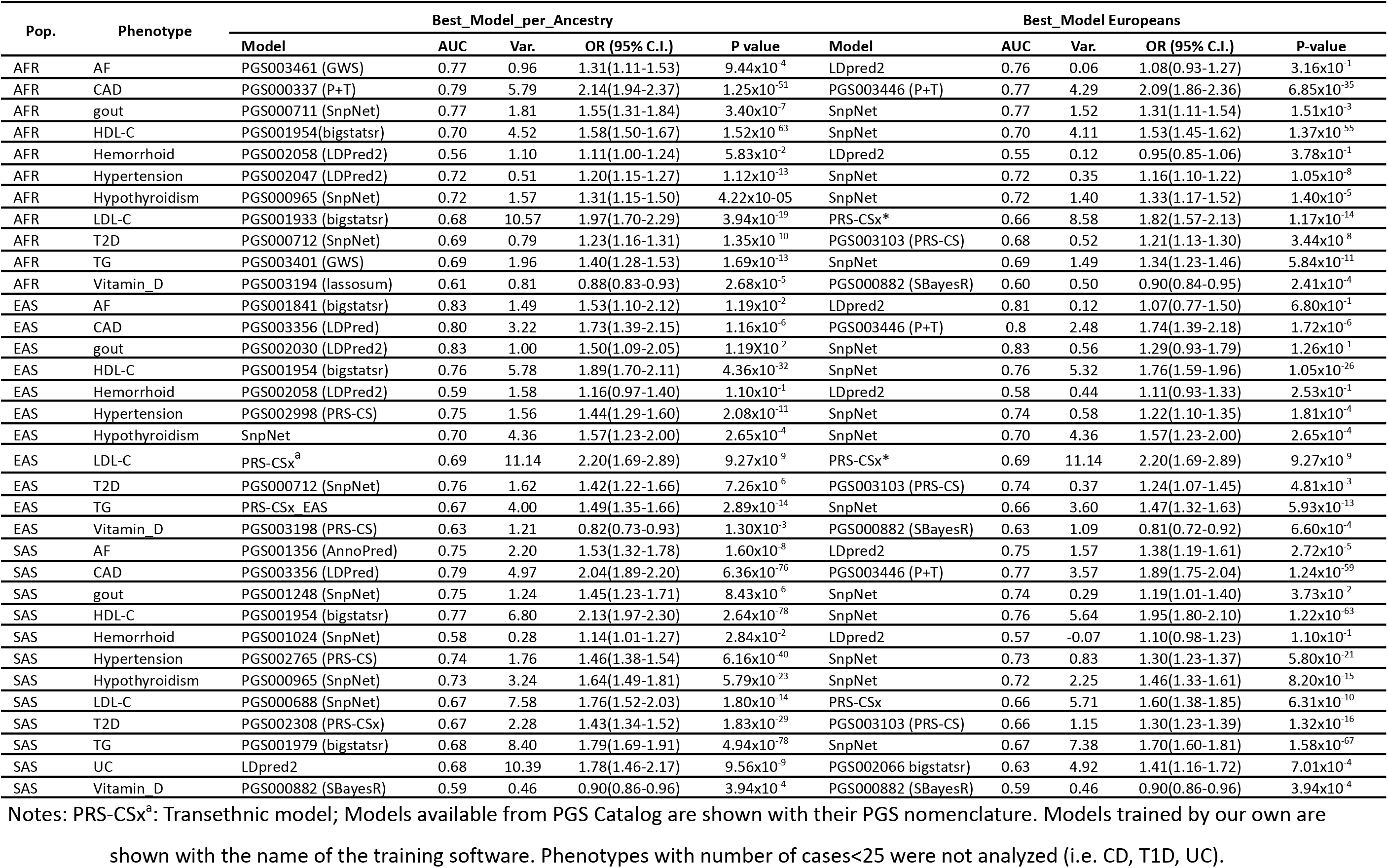
Comparison of best-performing PRS per ancestry for 14 medical conditions in non-European populations.

Our ancestry-specific selection increased the case enrichment in the top decile of the PRS distributions in non-European populations. Conditions improved the most from this approach includes hypertension in African individuals, HDL-C, LDL-C and T2D in East Asians, and CAD, hypothyroidism, and LDL-C in South Asians (Figure 3). A significant two-fold increased risk was observed between the 90^th^ and the intermediate percentiles for HDL levels (>60mg/dL) across all ancestries (i.e., OR(CI95) P value - AFR_HDL_ = 2.20 (1.82–2.67) 9.48 x 10^-16^; SAS_HDL_ = 3.33 (2.61–4.25) 6.13 x 10^-22^, EAS_HDL_ = 2.50 (1.71–3.68) 2.67 x 10^-6^). Those associations were also observed for CAD, high LDL levels (>190mg/dL) in African, and South Asian groups (i.e., OR(CI95) P value. AFR_CAD_ = 4.80 (3.52–6.61) 1.78 x 10^-22^; SAS_CAD_ = 3.92 (3.08–5.00) 2.16 x 10^-28^; AFR_LDL_ = 2.20 (1.46–3.30) 1.53 x 10^-04^; SAS_LDL_ = 2.65 (1.69–4.20) 2.38 x 10^-5^); (See Supplementary Table 9). Similar pattern was found for T2D in East Asians, and high TG levels (>200mg/dL) and hypothyroidism in South Asian ancestry. PRS percentile effects across different ancestries are shown in Supplementary Table 9 and Supplementary Figure 5.

**Figure 3.**
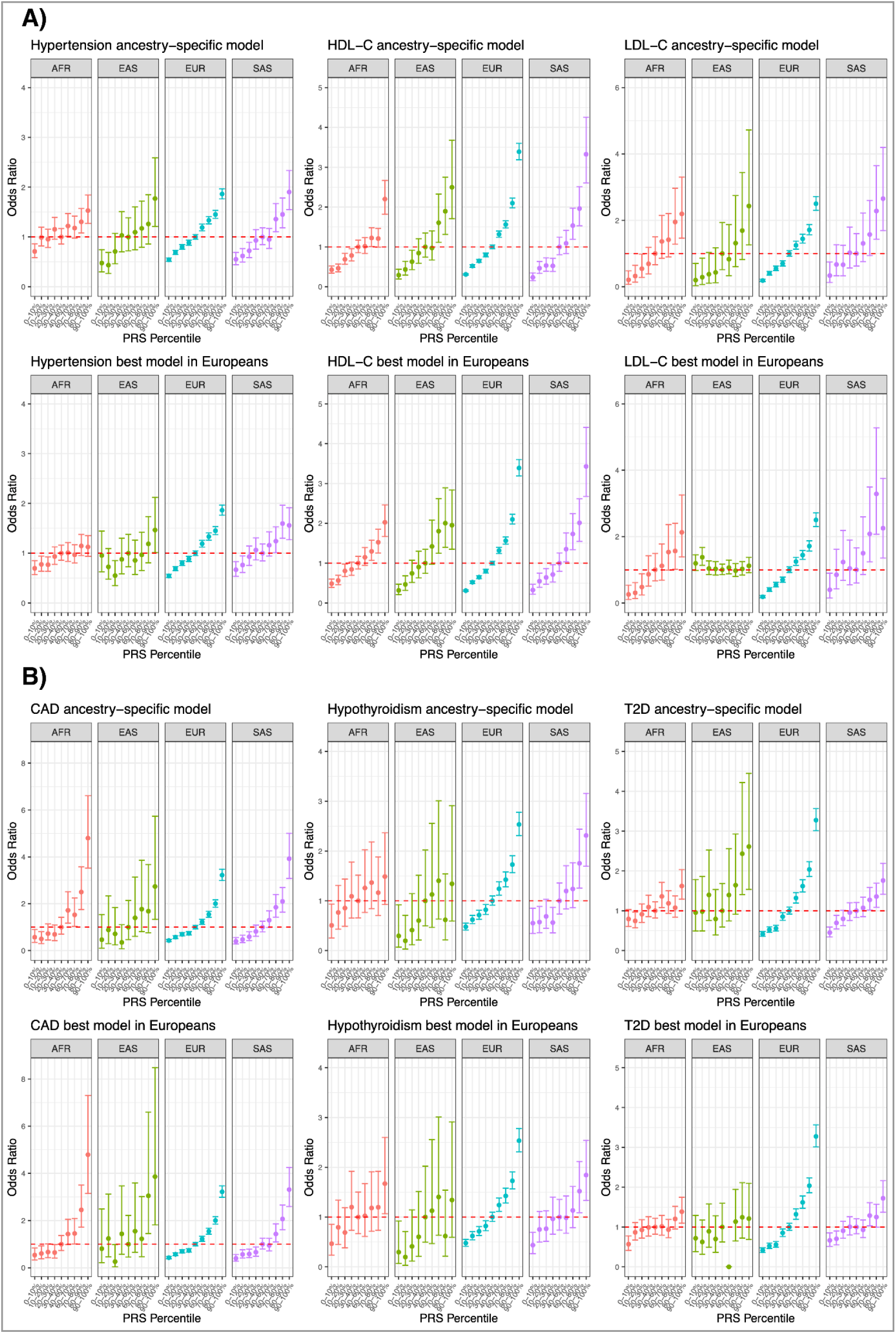
Effect of PRS percentile for hypertension, HDL-C, LDL-C, CAD, hypothyroidism, T2D across diverse ancestries. Red dashed line indicates OR=1.

### PRS-PheWAS across Complex Phenotypes

To assess whether our selected PRSs were associated with other medically relevant phenotypes, we performed a PRS-PheWAS across the conditions explored in this study in UK Biobank. We found 65 phenotype associations (43 and 22 for increasing and decreasing disease risk, respectively) across 14 medically relevant phenotypes (Figure 4, and Supplementary Table 11).

**Figure 4.**
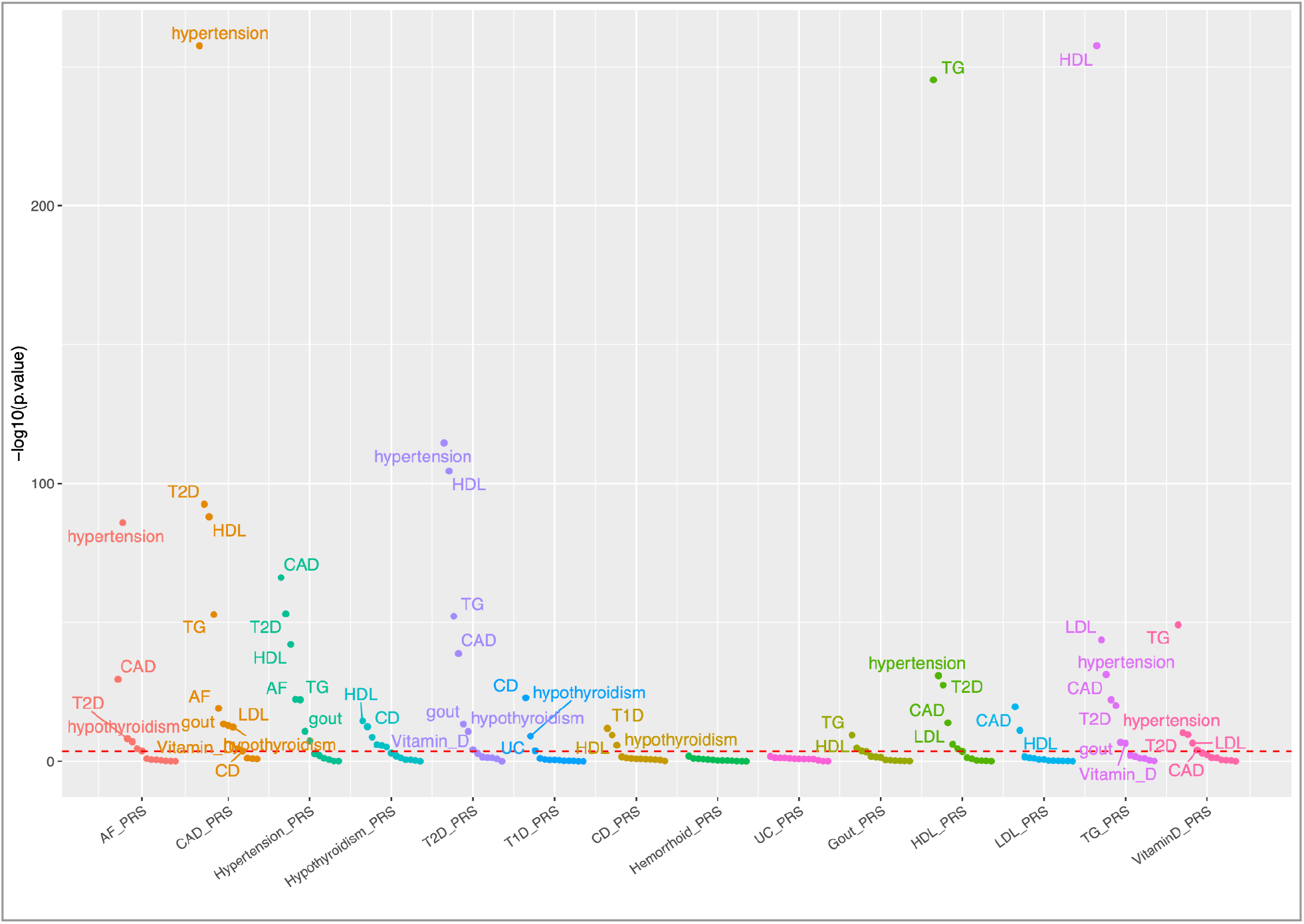
PRS-phenome wide association for fourteen medical conditions in European indviduals. Phenotypes are grouped according to the testing PRS. Red dashed line indicates the Bonferroni correction (2.74 x 10^-4^).

In particular, we identified two groups of conditions with cross-trait interrelationships, those presenting a cardiometabolic or an autoimmune component. Polygenic risk for CAD was associated with traits related to well-known risk factors, i.e., hypertension, altered lipid levels, and insulin resistance syndrome. PRSs for AF, CAD, hypertension, hypothyroidism, T2D and Lipid levels were highly associated with the risk of being diagnosed with other cardiometabolic conditions, highlighting the biological interplay among them. We did not find an association between T1D-PRS and the risk of suffering T2D, or the opposite (Supplementary Table 11). T1D-PRS was associated with CD, hypothyroidism, and UC, i.e., diseases well-known to present an autoimmune component. Low vitamin D PRS was significantly associated with the risk of developing conditions from the cardiometabolic axis, i.e., CAD, hypertension, T2D, and high levels of LDL and TG. These trends were similar to those observed for African, East Asian, and South Asian ancestry individuals (Supplementary Table 12), although most of the associations did not reach statistical significance, due to the decreased sample size (and thus decreased power) in non-European populations.

## 4. Discussion

In this study, we have conducted a wide assessment of PRS portability and performance across different ancestry groups, and we have contributed to independently validate PRS models. Our data supports the poor portability of European derived models to other ancestries, which is in line with prior findings [1][3][4][5][6]. Conversely, PRS performances relative to Europeans improved when we specifically selected the optimal PRS for a given condition and a given population ancestry, reinforcing the idea that optimal PRS methods are trait and context specific [11].

Our exploration of the prediction accuracy of PRS models in non-European ancestry cohorts has let us identify specific conditions (i.e., CAD, hypertension, hypothyroidism, HDL-C, LDL-C levels and T2D) where the application of PRSs in diverse populations showed more comparable performance to those observed in European groups. Specifically, we found that the PRS scores were more predictive when the selection of the appropriate PRS model for each particular individual was ancestry specific, which necessarily requires ancestry estimation prior to PRS estimation. It should be noted that the top selected models for CAD [22][23], were generated with genome-wide summary statistics derived from transethnic studies, which agrees with studies demonstrating the increased PRS predictability in multi-ethnic samples [11]. Although our approach does not solve the transferability problem impacting PRSs, we provide an alternative strategy to identify those conditions and ancestries where PRSs could work efficiently.

It should be noted that the gold standard solution to solve portability problems involves increasing the diversity of participants included and analyzed in genetic studies [10][24]. Training of new PRS models beyond European ancestries might lead to improved accuracy in genetic effect estimation particularly for variants with higher frequencies in non-European populations. It might also reduce the genetic distance from target and training samples, promoting PRS portability. In parallel, the generation of a new era of methods able to integrate ancestry deconvolution in PRS estimations might also support the generalization of PRS models.

We acknowledge some limitations and future improvements in our strategy. First, we observed disparities in the PRS effect sizes between UKBB and BbofA-GL cohorts for some conditions. In these studies, phenotypes were obtained using different phenotyping strategies - while ICD10 codes were extracted from UKBB, BBofA-GL administered self-reported questionnaires. Although the use of self-reported questionnaires does not offer the same level of granularity as clinical grade information, it is quickly becoming a common ascertainment strategy. In fact, direct-to-consumer genetic companies, which usually use this strategy, collectively account for millions of research participants and actively contribute to genetic studies [25]. Second, in this study we did not train and evaluate PRS predictability in admixed individuals. Genomes from admixed individuals consist of haplotypes from more than one ancestry group. It represents an extra-layer of complexity and presents technical limitations in the PRS field, despite offering the opportunity to yield novel genetic findings [6][26]. Most PRS methods depend on LD reference panels, and computational complexities in representing LD for admixed individuals hamper PRS modeling. Third, the generation of PRS models working for different populations is limited by the availability of combining multiple cohorts with diverse ancestries. Future studies will benefit greatly from the inclusion of more diverse datasets.

## 5. Conclusions

Overall, this study comprehensively evaluates PRS accuracy across a wide range of phenotypes for continental ancestries. Large variability of PRS performances across ancestral groups was observed when the evaluation was conducted using the best-PRS model for Europeans. We observed an improvement in the PRS performances when specifically selecting ancestry specific PRS models, reducing the uncertainty with respect to which PRS model to choose and in which ancestry group, and highlighting the benefit of incorporating ancestry inference in routine PRS evaluation. This approach has let us identify specific conditions with potential to be applied to different ancestries. Our work enables future studies to establish the best practices to evaluate PRS models, democratize the implementation of PRS models across populations and their application to translational research.

## Declarations

### Ethics approval and consent to participate

This study was approved by the institutional review board (IRB) at WCG IRB (https://www.wcgirb.com/) under IRB tracking number protocol number 20201332.

### Availability of data and materials

The BBofA-GL cohort that supports the findings of this study is available for qualified researchers at non-profit institutions upon entering into an agreement with Genomelink. All information will be shared subject to the above criterion upon request via.

### Authors’ contributions

SMG, MV, ALP designed the study. SMG, ALP, and DMM performed analysis of data. DMM, CQC, BM, MBT, MTML, ZY, KN, YM, JDW, AI, NK, TT, and CDB provided interpretation of the results. SMG, JDW NK and CDB drafted the manuscript. All authors contributed critically, read, revised, and approved the final version.

## Supporting information

Supplementary Figure 1. Principal component analysis for genetic ancestry in the UKBB.

Supplementary Figure 2. Principal component analysis for genetic ancestry in the BbofA-GL.

Supplementary Figure 3. Effect of PRS percentile for 14 medical conditions

Supplementary Figure 4. Percentage of variance explained by the PRS relative to European-ancestry individuals

Supplementary Figure 5a. Effect of PRS percentile for AF, Gout, and hemorrhoid across diverse populations

Supplementary Figure 5b. Effect of PRS percentile for UC, TG and Vitamin D levels across diverse populations

## Acknowledgements

The authors would like to thank all the participants for being a part of this study.

