## Supplementary Figure 1. Principal component analysis for genetic ancestry in the UKBB. for "Polygenic risk score portability for common diseases across genetically diverse populations"

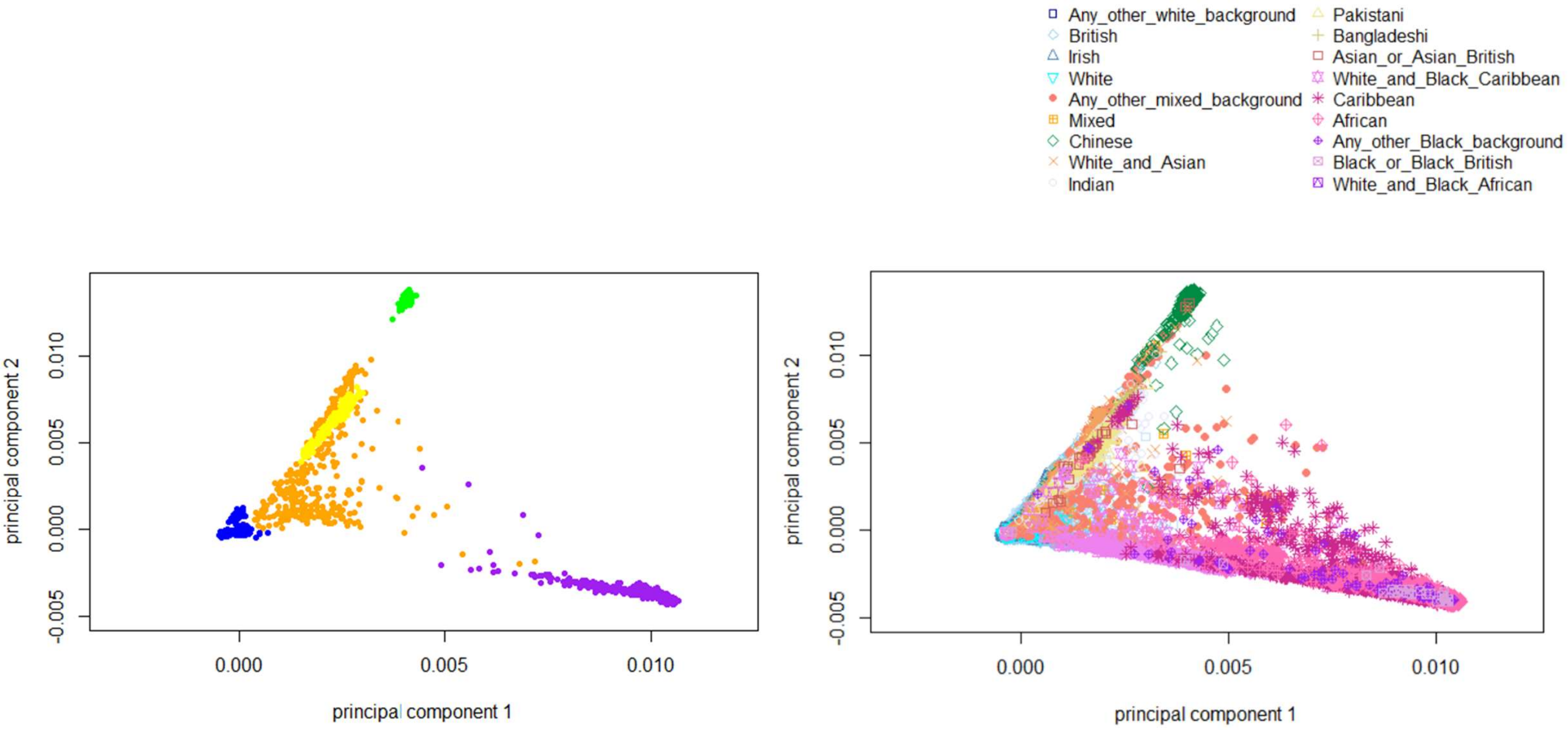


a. Reference population 1000G

b. UKBB Dataset (406K)

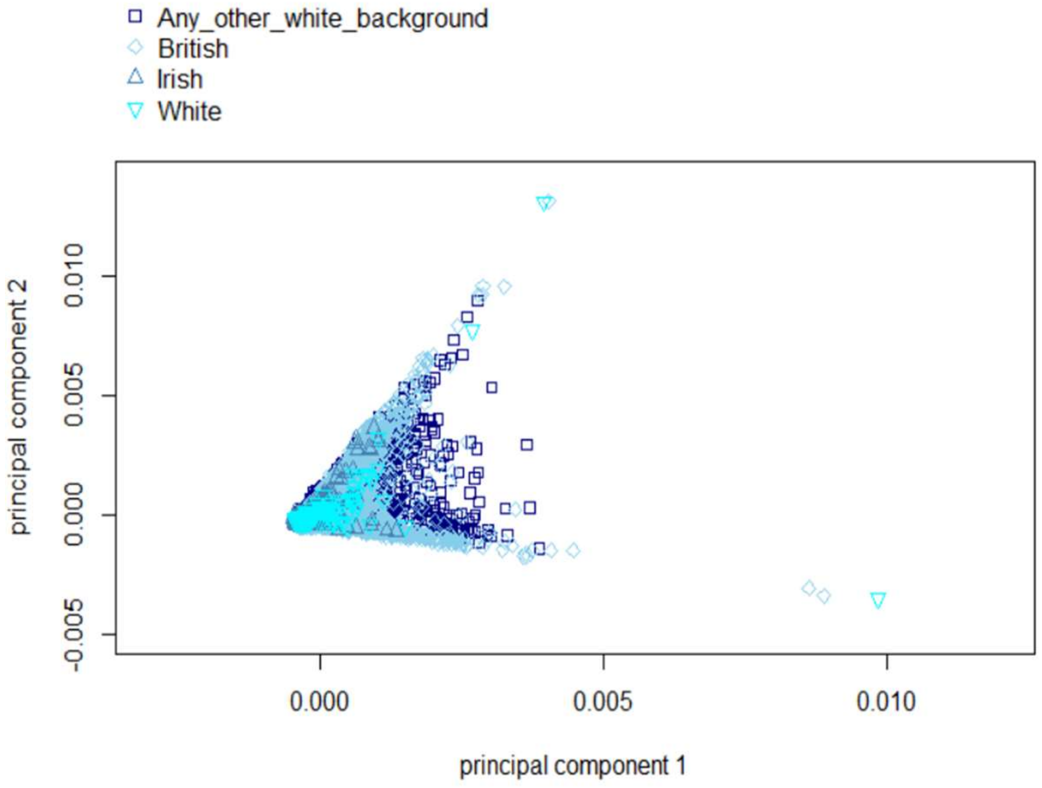


c. UKBB Dataset – Individuals self-identified as White background

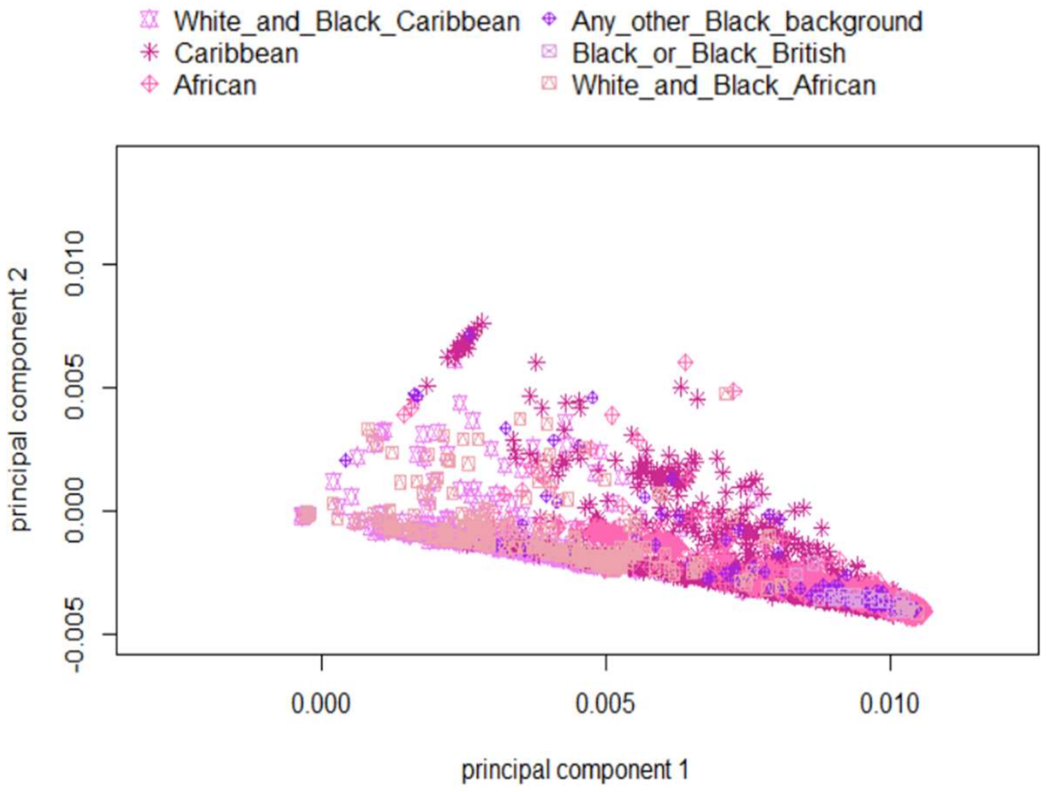


d. UKBB Dataset – Individuals self-identified as African background

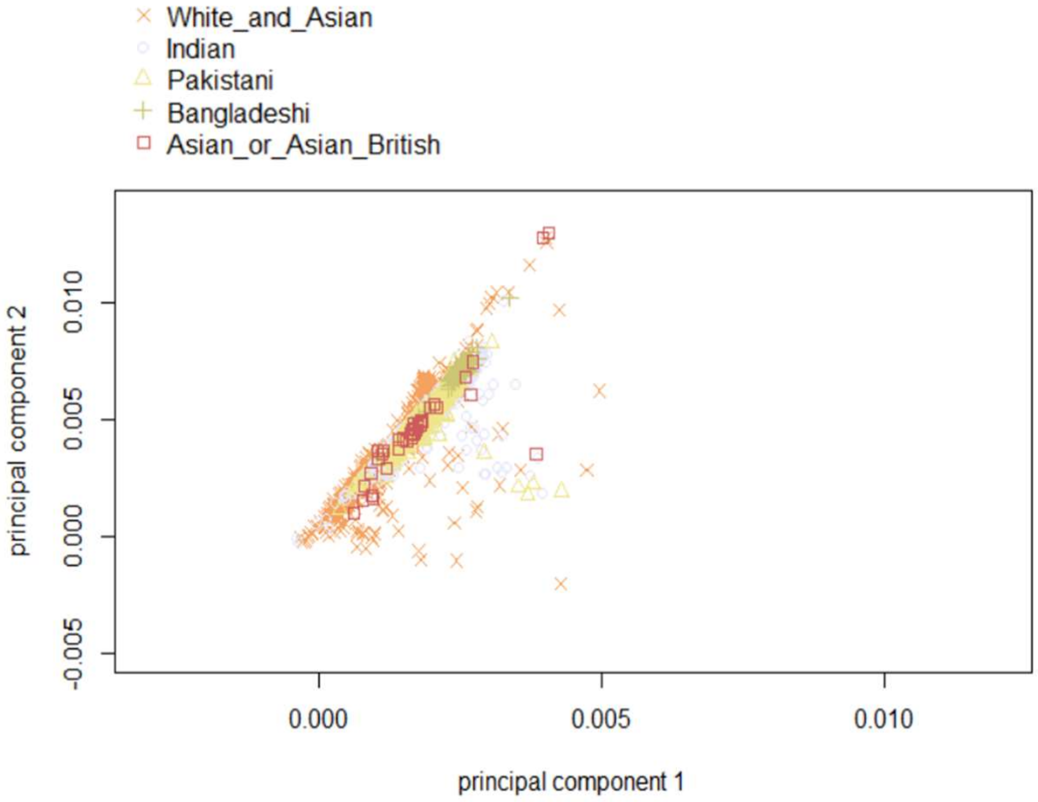


e. UKBB Dataset – Individuals self-identified as South Asian background

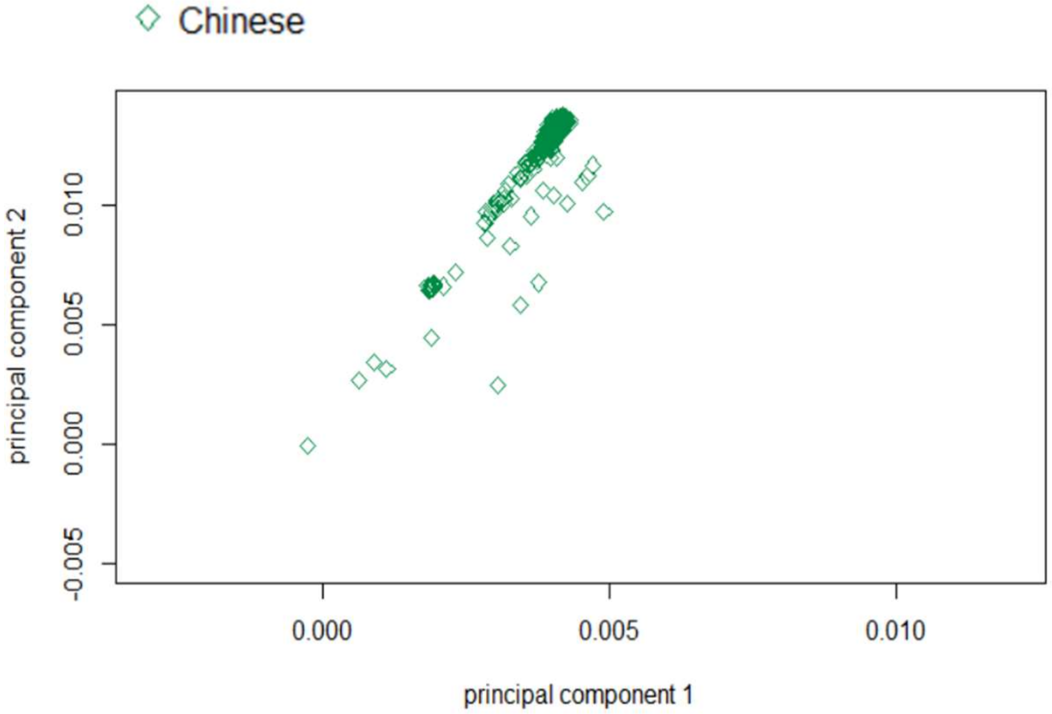


f. UKBB Dataset – Individuals self-identified as East Asian background
