## Supplementary figures and images for "Polygenic risk score portability for common diseases across genetically diverse populations"

### Supplementary Figure 2. Principal component analysis for genetic ancestry in the BbofA-GL.

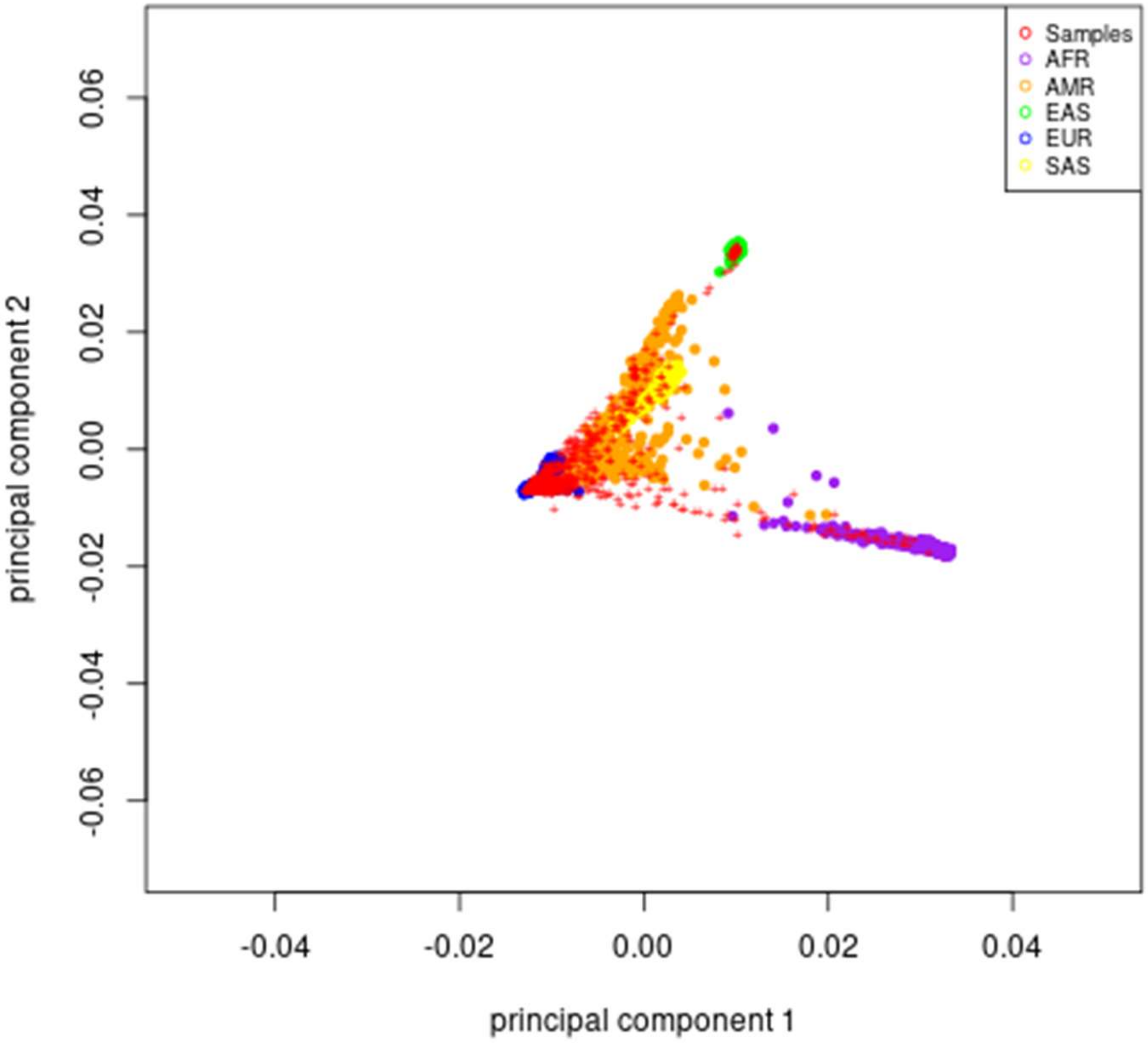

### Supplementary Figure 3. Effect of PRS percentile for 14 medical conditions

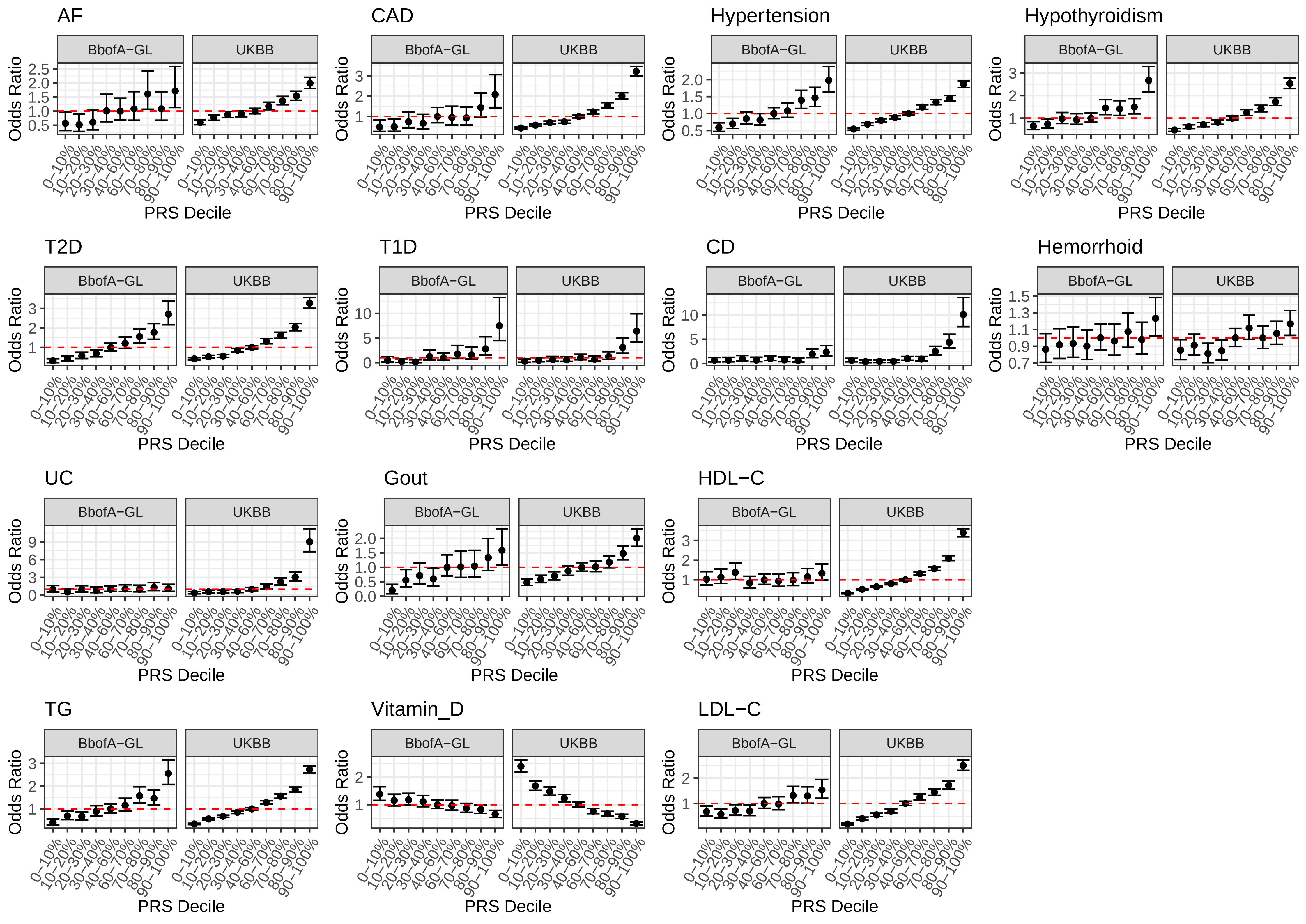

### Supplementary Figure 4. Percentage of variance explained by the PRS relative to European-ancestry individuals

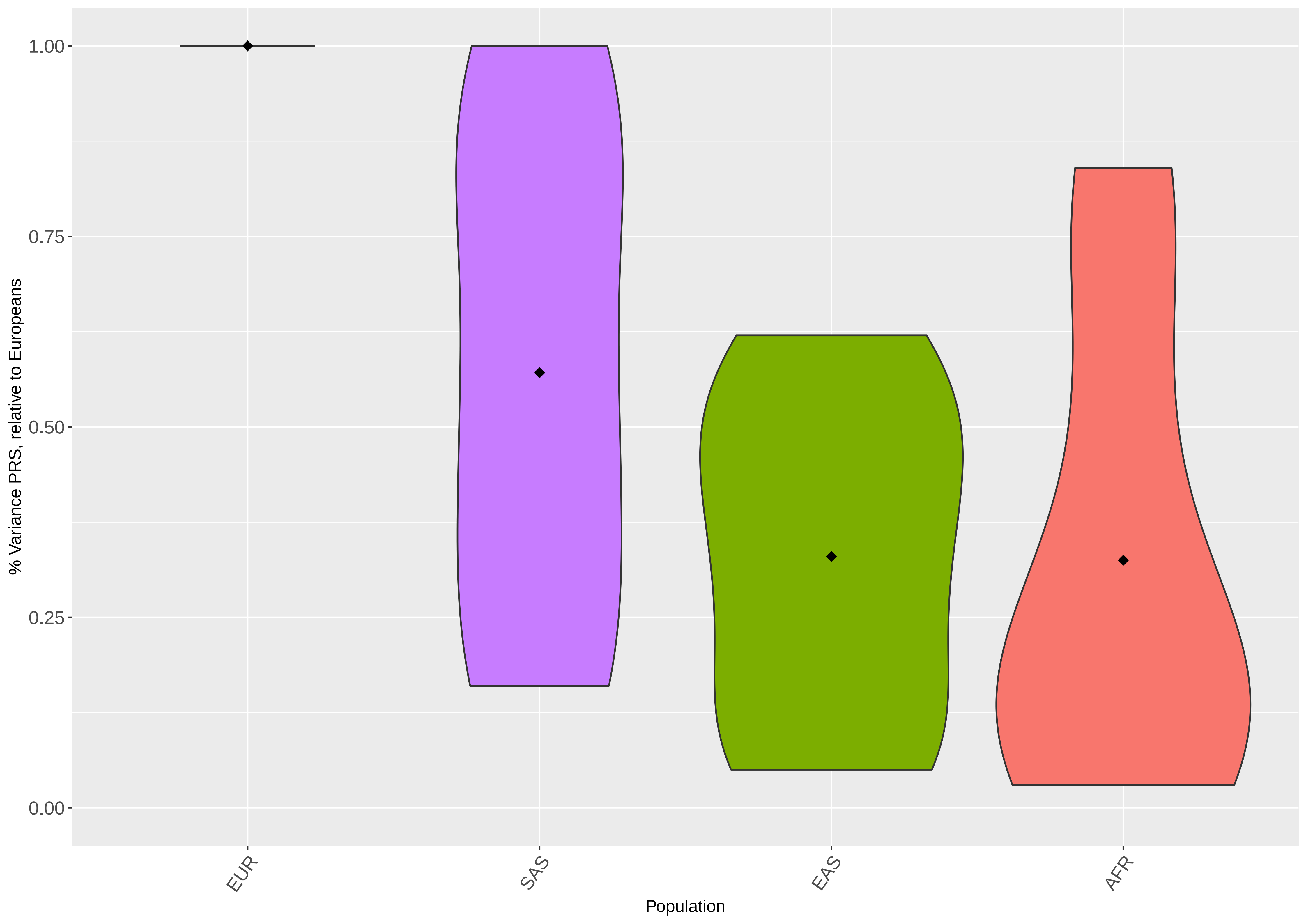

### Supplementary Figure 5a. Effect of PRS percentile for AF, Gout, and hemorrhoid across diverse populations

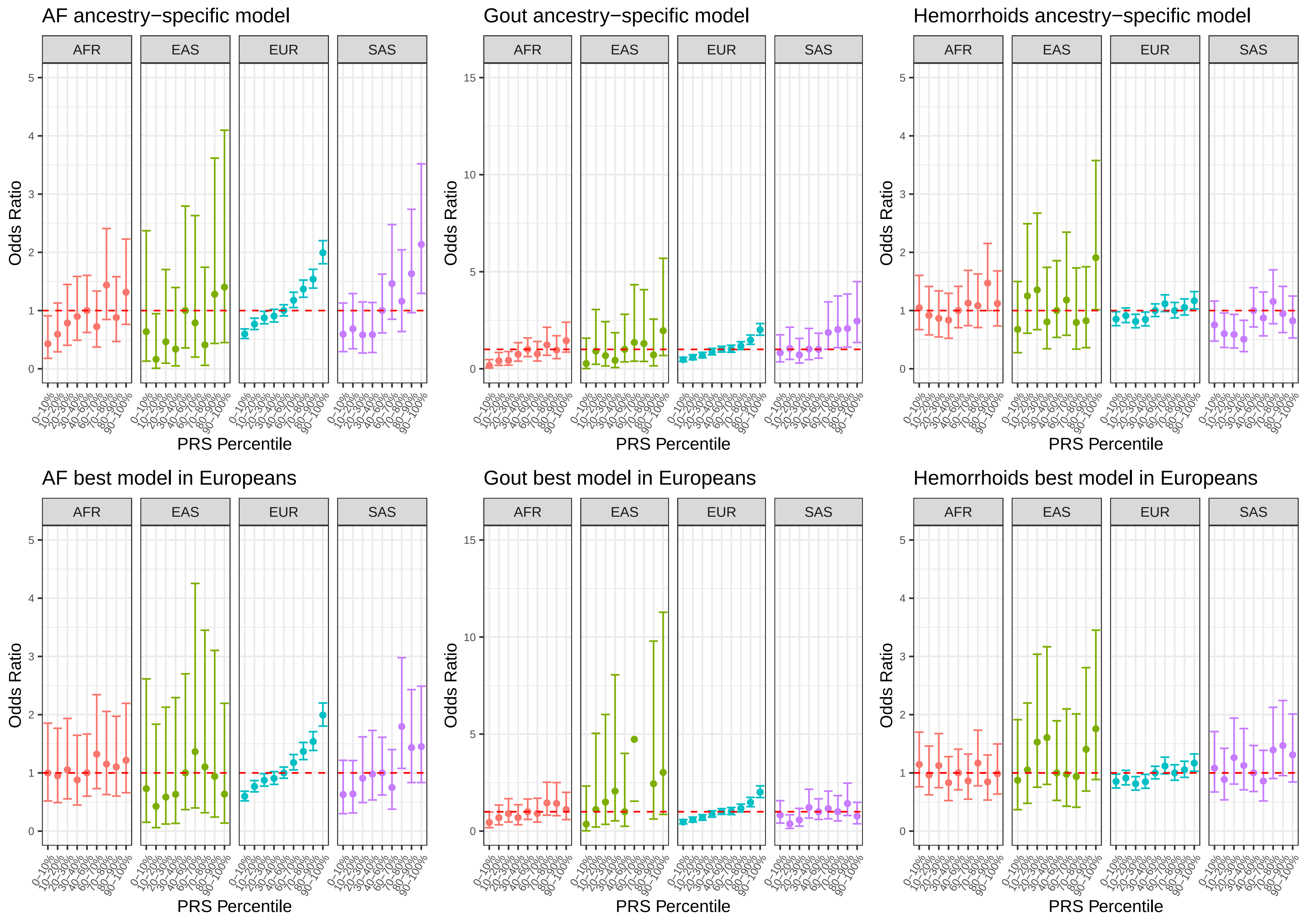

### Supplementary Figure 5b. Effect of PRS percentile for UC, TG and Vitamin D levels across diverse populations

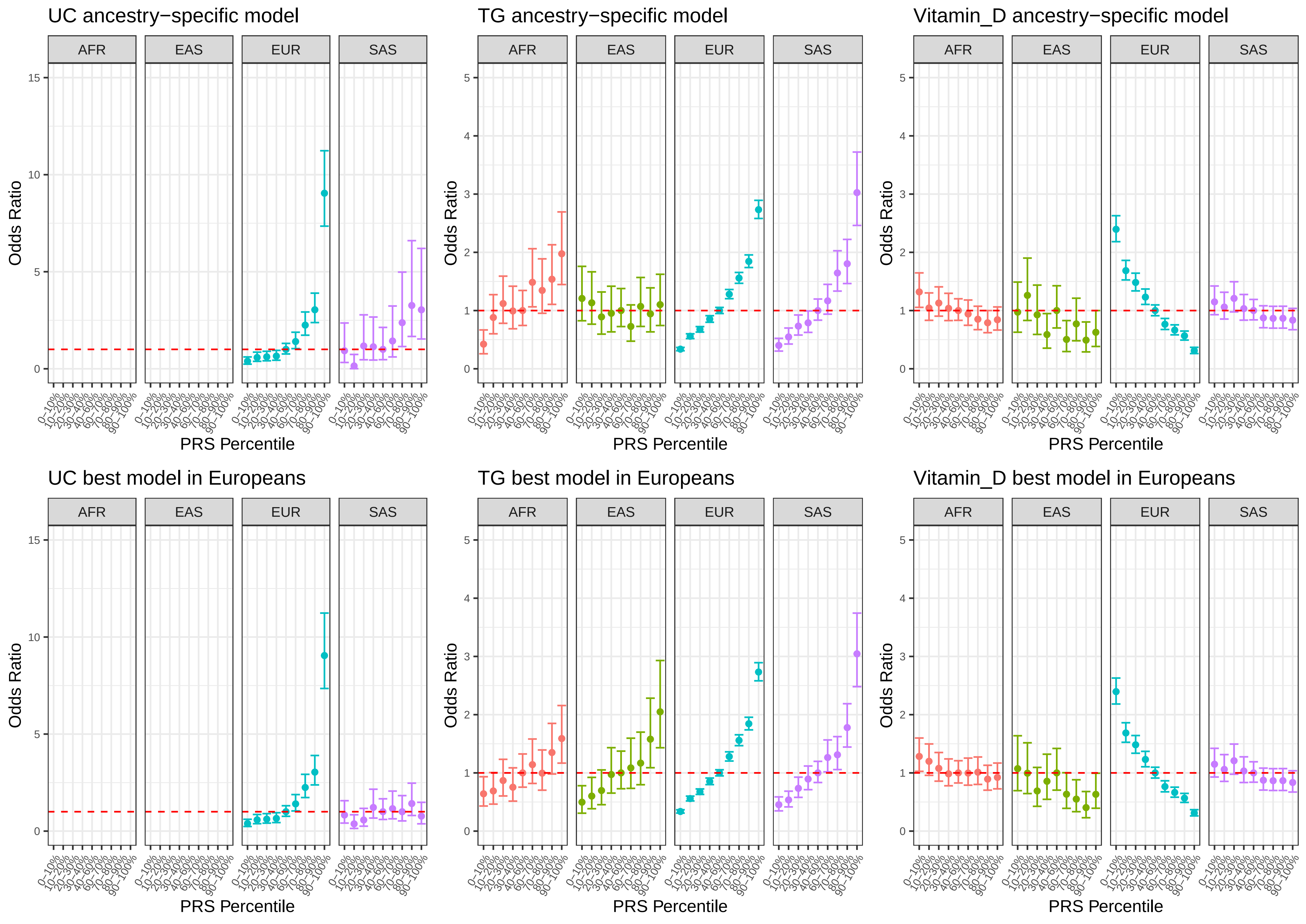
